# DBS-induced gamma entrainment as a new biomarker for motor improvement with neuromodulation

**DOI:** 10.1101/2024.04.25.24306357

**Authors:** Varvara Mathiopoulou, Jeroen Habets, Lucia K. Feldmann, Johannes L. Busch, Jan Roediger, Jennifer K. Behnke, Gerd-Helge Schneider, Katharina Faust, Andrea A. Kühn

**Affiliations:** Department of Neurology, Charité—Universitätsmedizin Berlin, corporate member of Freie Universität Berlin and Humboldt-Universität zu Berlin, Berlin, Germany; Berlin Institute of Health at Charité Universitätsmedizin Berlin, Berlin, Germany; Department of Neurosurgery, Charité—Universitätsmedizin Berlin, corporate member of Freie Universität Berlin and Humboldt-Universität zu Berlin, Berlin, Germany; Berlin School of Mind and Brain, Charité Universitätsmedizin Medicine, Berlin, Germany; NeuroCure Clinical Research Centre, Charité Universitätsmedizin, Berlin, Germany; DZNE, German Center for Degenerative Diseases, Berlin, Germany

## Abstract

Finely tuned gamma oscillations have been recorded from the subthalamic nucleus and cortex in Parkinson’s disease patients undergoing deep brain stimulation and are often associated with dyskinesia. More recently, it was shown that deep brain stimulation entrains finely tuned gamma to ½ of the stimulation frequency; however, the functional role of this signal is not yet fully understood.

We recorded local field potentials from the subthalamic nucleus in 19 chronically implanted Parkinson’s disease patients under effective dopaminergic medication and during deep brain stimulation with increasing stimulation amplitude, while they were at rest and during repetitive hand movements. We analyzed the effect of stimulation intensity on gamma band 1:2 entrainment and compared the entrained signal during rest and during repetitive movement.

Spontaneous finely tuned gamma was present in eight out of 19 patients (peak frequency *μ* = 78.4 ±4.3 Hz). High-frequency deep brain stimulation induced 1:2 gamma entrainment in 15 out of 19 patients. Entrainment occurred at a mean stimulation amplitude of 2.2 0.75 mA and disappeared or decreased in power during higher stimulation amplitude in three patients. In patients with spontaneous finely tuned gamma, increasing the stimulation amplitude induced a progressive frequency shift of spontaneous finely tuned gamma until it locked to 1:2 entrainment. Only five out of 15 patients with entrained gamma activity showed dyskinesia during stimulation. Further, there was a significant increase in the power of 1:2 entrained gamma activity during movement in comparison to rest. Finally, patients with entrained gamma activity had faster movements as compared to those without gamma entrainment.

These findings argue for a functional relevance of the stimulation-induced 1:2 gamma entrainment in Parkinson’s disease patients as a prokinetic activity that, however, is not necessarily promoting dyskinesia. Previously published electrophysiological models of entrainment fit well to our results and support our findings that stimulation-induced entrainment can be a promising real-life biomarker for closed-loop deep brain stimulation.

## Introduction

Deep brain stimulation (DBS) is an established treatment option for patients with Parkinson’s disease, especially those with pronounced motor fluctuations.^1^ Implantable pulse generators (IPG) deliver electrical stimulation to deep subcortical structures, e.g., the subthalamic nucleus (STN) or the globus pallidus internus. The latest generation of commercially available IPGs enable the recording of local field potentials (LFPs) from these target regions during active stimulation in chronically implanted patients.^2^ This novel setup allows for the chronic evaluation of electrophysiological biomarkers, such as beta band activity (13-35 Hz) that has been explored in acute settings over the last decades, to develop feedback signals for closed-loop DBS.

Within this framework, pathologically enhanced beta band has been related to bradykinesia and rigidity in PD and is considered an antikinetic activity, whereas gamma band has been linked to movement and dyskinesia as a prokinetic activity.^3^ Oscillatory gamma band plays a pivotal role in orchestrating large-scale network activity in a variety of cognitive and motor phenomena.^4^ In the realm of movement, a broader gamma band (40-90 Hz) has been closely linked to the initiation and execution of movements and is considered to have a prokinetic effect.^5^ In PD patients, a more narrow frequency band (70-90 Hz), referred to as spontaneous finely tuned gamma (FTG), is associated with levodopa-induced dyskinesia (LID), a condition in which PD patients experience involuntary movements as a result of long-term treatment with dopaminergic medication.^6–10^ Recently, LFPs recorded during high-frequency DBS in patients with PD unveiled an entrained neural signal within the gamma band (40-100 Hz) in both cortical and subcortical recordings.^6,11,12^ Neural entrainment describes the phenomenon by which ongoing brain oscillations synchronize to the rhythmic pattern of an external stimulus, such as repetitive auditory cues, visual signals, or electrical stimulation.^13–17^ Successful neural entrainment by an external stimulus enhanced the cognitive or motor function that was associated to the oscillatory activity being entrained.^18,19^ In patients with PD, DBS is typically applied in frequencies between 90 and 180 Hz. DBS-induced entrainment was identified in ½ of the DBS frequency, and was reported in several clinical conditions, i.e., with and without dopaminergic medication or dyskinesia.^9,12^ Furthermore, 1:2 entrainment is hypothesized to appear only in certain combinations of DBS amplitude and frequency and differs between individuals.^20,21^

Given the role of gamma band activity in a variety of motor phenomena, it is important to understand how DBS modulates ongoing gamma band oscillations, and what is the functional relevance of a potentially entrained signal.^8,12^ Here, we study 1:2 entrainment in the gamma band in subthalamic LFP recordings of chronically implanted PD patients under simultaneous dopaminergic treatment and high-frequency DBS. First, we show that both DBS amplitude and frequency modulated gamma entrainment. Second, spontaneous FTG activity, when present, gradually entrained to the ½ DBS frequency with increased DBS intensity. Last, we highlight the functional relevance of this entrained signal, by showing that repetitive finger tapping increased or induced gamma entrainment and that patients with gamma entrainment had a better motor performance.

## Materials and methods

### Subjects and study protocol

We included 19 PD patients (10 female, mean age = 62.6 years ±6.7, mean disease duration = 10.9 ±3.7 years, 19 STNs) treated with STN DBS who were enrolled during standardized 3 or 12-month follow up visits at the Department of Movement Disorders and Neuromodulation. The placement of the electrodes was determined by stereotactic planning, intraoperative microelectrode recordings and clinical testing, and was confirmed by post-operative imaging.^22^ The patients were implanted with Medtronic 3389 (*n* = 9) or SenSight (*n* = 10) DBS electrodes, which were connected to the sensing enabled ‘Percept’ IPG (Medtronic ©, Minneapolis, MN, USA). Demographic and clinical details are presented in **Table 1**.

**Table 1.** Demographic and clinical details.

|  | PATIENT | AGE RANGE (Y) | SEX | DD (Y) | FOLLOW-UP (MONTHS) | LEDD (MG) | ELECTRODE | STIM CONTACT | RECORDING CONTACTS | CONTACTS FOR CHRONIC STIMULATION | GAMMA PEAK DBS-OFF (HZ) | ENTRAINMENT ONSET (MA) | UPDRS MED OFF-STIM OFF/MED ON-STIM OFF | DYSKINESIA DBS OFF/ON |
| --- | --- | --- | --- | --- | --- | --- | --- | --- | --- | --- | --- | --- | --- | --- |
| WITH FTG DBS -OFF | #1 | 60-65 | f | 10 | 3 | 775 | 3389 | L2 | L1-3 | 1 | 77 | 1.0 | 22/5 | No/No |
|  | #2 | 55-60 | f | 13 | 6 | 300 | 3389 | L1 | L0-2 | 1 | 81 | 2.0 | 78/34 | Mild/Mild |
|  | #3 | 50-55 | m | 7 | 3 | 500 | 3389 | R1 | R0-2 | 1 | 73 | 2.5 | 51/28 | No/No |
|  | #4 | 65-70 | m | 15 | 3 | 300 | 3389 | R2 | R1-3 | 1 | 73 | 2.5 | 39/11 | Mild/Moderate |
|  | #5 | 70-75 | f | 12 | 12 | 500 | SenSight | R2 | R1-3 | 3 | 76 | 2.4 | 44/17 | No/Mild |
|  | #6 | 50-55 | m | 13 | 12 | 0 | SenSight | L2 | L1-3 | 1,3 | 84 | 1.0 | 43/30 | No/Moderate |
|  | #7 | 50-55 | m | 14 | 3 | 535 | SenSight | L1 | L0-2 | 1 | 80 | 2.5 | 19/15 | No/No |
|  | #8 | 65-70 | f | 7 | 18 | 275 | SenSight | R2a | R1-3 | 1a,2a | 83 | 1.4 | 53/43 | Moderate/Severe |
| NO FTG DBS-OFF | #9 | 65-70 | m | 11 | 12 | 1250 | 3389 | L2 | L1-3 | 2 | (-) | 2.0 | 54/44 | No/No |
|  | #10 | 70-75 | m | 20 | 12 | 1000 | SenSight | R2b | R1-3 | 2 | (-) | 2.8 | 49/39 | No/No |
|  | #11 | 70-75 | f | 7 | 12 | 475 | SenSight | R1 | R0-2 | 1a,1c | (-) | 1.5 | 29/19 | No/No |
|  | #12 | 70-75 | f | 14 | 12 | 375 | SenSight | R1b | R0-2 | 0,1 | (-) | 3.0 | 51/39 | No/No |
|  | #13 | 55-60 | m | 15 | 12 | 550 | SenSight | R1c | R0-2 | 1 | (-) | 2.0 | 34/27 | No/No |
|  | #14 | 55-60 | m | 7 | 12 | 350 | SenSight | R2a | R1-3 | 2a,2b,3 | (-) | 3.7 | 57/35 | No/No |
|  | #15 | 66-70 | f | 7 | 12 | 400 | SenSight | L2b | L1-3 | 2b,2c | (-) | 2.5 | 42/35 | No/No |
| NO GAMMA | #16 | 60-65 | f | 7 | 3 | 525 | 3389 | L2 | L1-3 | 2 | (-) | (-) | 42/28 | No/No |
|  | #17 | 60-65 | f | 7 | 12 | 0 | 3389 | R2 | R1-3 | 2,3 | (-) | (-) | 40/28 | No/No |
|  | #18 | 55-60 | m | 11 | 12 | 200 | 3389 | R1 | R0-2 | 2 | (-) | (-) | 38/- | No/No |
|  | #19 | 65-70 | f | 11 | 12 | 1000 | 3389 | R1 | R0-2 | 0,1 | (-) | (-) | 67/60 | No/No |

DD = Disease duration; LEDD = Levodopa equivalent daily-dose

All recordings were performed ON medication 30 min after patients received 100-200 mg fast-acting L-Dopa. LFPs were recorded during a systematic unilateral ramping of stimulation amplitude at rest and during three runs of 10 seconds repetitive finger-tapping. DBS pulse width and frequency were set at 60 µs and 125 Hz (SenSight electrodes) or 130 Hz (3389 electrodes). Patients performed all recordings seated comfortably in an armchair. All sessions were video recorded. LFPs were recorded in a bipolar montage between the two contact-rings adjacent to the stimulation level, according to standard Medtronic Percept settings.^2^ Nine patients (all with 3389 circular electrodes) were reported previously.^23,24^ In these cases contact selection was based on the strongest beta activity (13-35 Hz) during OFF-medication. Contact selection in the remaining 10 patients (SenSight electrodes) was based on the occurrence of gamma entrainment during a monopolar review in OFF-medication, which is described elsewhere.^25^ Stimulation was applied at single segments (*n* = 6) or at the ring level (*n* = 4). Additionally, in five of those patients (patients #8, 10, 12, 13, 15) the protocol was repeated at three different DBS frequencies (i.e., 110, 125, and 145 Hz).

All patients gave written informed consent prior to the study. The study protocol was approved by the local ethics committee (Medical Ethical Committee of Charité Universitätsmedizin, Berlin, Germany, Protocol ID.: EA2/256/60) in accordance with the standards set by the Declaration of Helsinki.

### Data Acquisition and Analysis

#### LFP recordings and analysis

Continuous time series of LFPs originating from the STN were recorded using the BrainSense Streaming feature of the Percept IPG with a sampling frequency of 250 Hz. LFPs were preprocessed using the open source ‘PERCEIVE Toolbox’ and further analysis was performed using Python.^26,27^ Data were transformed to the time-frequency domain using a Fast Fourier Transform with a window size of one second and a 25% overlap. Rest activity was isolated during all stimulation conditions for each patient. Gamma peak frequencies were defined as the frequency with the largest amplitude between 40 and 100 Hz. Peaks of gamma in FTG/entrainment were selected visually and were the only distinct peaks with the highest amplitude in 40-90 Hz.

We further investigated the effects of varying DBS settings on three different gamma phenomena, i.e., FTG without stimulation (referred to as ‘spontaneous FTG’), 1:2 entrainment, and the intermediate activity between these two. To obtain these three signals, raw data were filtered and averaged 5 Hz around the subject-specific spontaneous FTG peak (e.g., 78 Hz: 76–80 Hz), 5 Hz around the subharmonic frequency at ½ the DBS frequency (e.g., 65 Hz: 63– 67 Hz), and between the spontaneous-FTG and entrainment (i.e., in this example 67–76 Hz). The analytic signal was calculated with the Hilbert transform. Next, we took the absolute analytical signal, z-scored it to allow for an inter-subject comparison, and smoothed it with a moving average of 2 seconds. For the analysis, we divided the recordings of subjects with entrainment into three phases: DBS-OFF, DBS-ON before the entrainment onset, and DBS-ON after the entrainment onset. Since these phases have varying durations, we resized each phase to a common duration by resampling the smoothed z-scored analytic signal. Resampling the z-scored analytical signals preserved the overall dynamics of the signal, while allowing for group level analyses.

### Motor assessment

Clinical improvement with dopaminergic medication was assessed as change in Unified Parkinson’s Disease Rating Scale Part III (UPDRS III) scores before and after dopaminergic medication intake. The presence and absence of dyskinesia was assessed by a clinician present at the recordings and assessed using the Clinical Dyskinesia Rating Scale.^28^ Change in motor performance during stimulation was assessed by finger tapping. To quantify the motor task, a tri-axial accelerometer was attached to the index finger opposite to the side where the unilateral DBS was applied. Accelerometer traces were captured at a sampling frequency of 4 kHz (TMSi Saga) and were down sampled to 250 Hz. The traces were synchronized to the LFPs using a custom-made synchronization device. This device produced a visually detectable signal in the video recordings, while simultaneously generating a detectable vibration in the accelerometer traces.

Movement blocks were considered during highest DBS amplitude with entrainment in the cohort in which entrainment was induced, and during the highest DBS amplitude applied in the cohort without entrainment. Movement traces were further processed and analyzed with the open-source algorithm ‘ReTap’.^29^ ReTap was used to automatically detect both single taps and ten-second blocks of finger tapping movement. Based on the performance and diversity of kinematic concepts captured, we included the following four movement metrics in our kinematic analysis: normalized root mean square, mean finger raise velocity (m/sec), coefficient of variation of intra-tap intervals, and coefficient of variation of root mean square impact. For further details we refer to the ReTap validation study.

### Statistical Analysis

Wilcoxon signed-rank test was used for group comparisons and Spearman rho for correlations. A t-test was used to compare the power of entrainment during movement in comparison to baseline. A linear mixed effect model was used to investigate the relationship between the presence of entrainment and the motor performance, while correcting for the repetition of movement blocks within patients with a random intercept.

### Data Availability

The data that support the findings of this study are available on request from the corresponding author. The data are not publicly available due to GDPR-compliance rules.

## Results

### 1. DBS-induced gamma entrainment is amplitude dependent and is modulated by the DBS frequency

Eight patients displayed spontaneous FTG activity ON medication without stimulation (53%, Patients #1-8, peak frequency *μ* = 78.4 ±4.3 Hz). All these patients showed gamma entrainment with stimulation. Additionally, seven patients without spontaneous FTG developed a 1:2 gamma entrainment with increase in stimulation amplitude during ipsilateral DBS. Thus, 15 out of 19 patients (79%) showed gamma entrainment at variable stimulation amplitude during DBS. Gamma entrainment in an example LFP recording is shown in **Supplementary Fig 1.** Peak frequency of entrained activity followed a strict 1:2 pattern (i.e. 62.5 Hz with 125 Hz stimulation frequency, 65 Hz with 130 Hz stimulation frequency). The entrainment occurred at a mean stimulation amplitude of 2.19 ±0.75 mA and disappeared when DBS was switched off (**Figure 1a)**. Additionally, the entrainment disappeared in two cases (#2 and #14) with higher stimulation intensities and was reduced in amplitude in one case (#1). The four remaining patients (21%) showed no 1:2 entrainment (#16-19, for an overview see **Figure 1b**).

**Figure 1.**
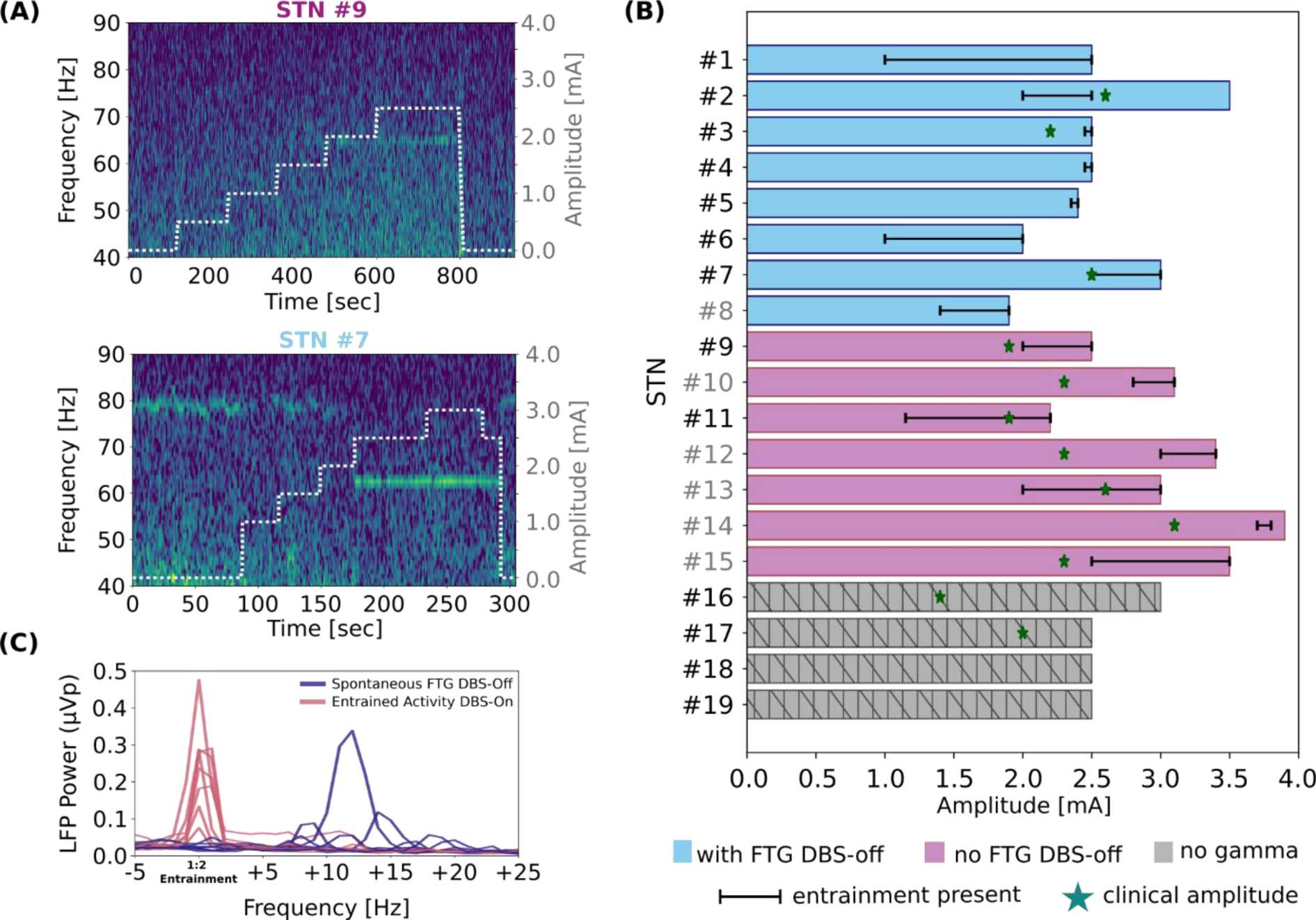
DBS-induced entrainment is modulated by the DBS amplitude. **(A)** Example cases of entrainment at 65 Hz induced by DBS in the absence (upper plot) or in the presence (lower plot) of spontaneous FTG activity during DBS-off. Entrainment appears with increased amplitude and disappears when DBS is switched off. When there is spontaneous FTG during DBS off (lower plot, here around 77 Hz), this adjusts to ½ of DBS frequency, and it resets when DBS is off again at the end of the recording. **(B)** Schematic presentation of all recordings included in the study. Bar plots along the x-axis show the highest amplitude that was used for every patient. Black lines show the amplitude range in which the entrainment was present. Recordings from STNs #1-8 showed spontaneous FTG in the absence of DBS. Recordings from STNs #9-15 did not show spontaneous FTG in the absence of DBS. Recordings from STNs #16-19 did not show any activity within the gamma band on/off DBS. Green stars show the amplitude that was selected during DBS optimization as clinically most beneficial in those cases where the clinically used electrode contact was the same as the one that was used for LFP recordings. Grey STN indices (e.g., #8) refer to the STNs that were stimulated directionally. **(C)** Mean power spectra of 30 sec of rest on/off DBS from all eight STNs in which there was spontaneous FTG during DBS-off. Peaks during entrainment have higher amplitude and are narrower in comparison to peaks during DBS-off.

The patients showing spontaneous FTG developed 1:2 entrainment from lower DBS amplitudes onwards compared to the patients without spontaneous FTG, although this difference did not reach statistical significance (group with spontaneous FTG *μ* = 1.91 ±0.68 mA, group without *μ* = 2.6 ±0.62 mA, *P* = 0.162). In the same group we compared peak amplitudes and widths of spontaneous FTG and 1:2 entrainment during rest. Peaks during entrainment had higher amplitudes (*P* = 0.049) and narrower widths (*P* < 0.001) than peaks during spontaneous FTG (**see Figure 1c**). There was no correlation between the power of entrainment and the difference between the spontaneous FTG peak and the entrainment peak frequencies (*P* = 0.365).

Regarding clinical features, three out of eight patients with spontaneous FTG exhibited dyskinesia ON medication without stimulation. Two additional patients from this group developed dyskinesia during stimulation and in two patients dyskinesia increased with stimulation. Patients without spontaneous FTG did not develop dyskinesia during stimulation despite gamma entrainment. UPDRS III improvement with levodopa challenge during recordings was larger in the group with spontaneous FTG (47.8%) as compared to those without (24.7%; *P* = 0.024). The DBS amplitudes that resulted in entrainment in each STN are shown in **Figure 1b**. Clinical details are shown in **Table 1**.

Last, we inspected the adjustment of 1:2 entrainment frequency when different DBS frequencies were applied (i.e., 110, 125, and 145 Hz) in five patients, in one hemisphere per patient. Four out of five patients showed 1:2 entrainment that adjusted to 55, 63, and 73 Hz respectively during all three DBS frequencies (**Figure 2a)**. One patient showed 1:2 entrainment only during DBS at 145 Hz. There was no significant difference in the stimulation amplitude needed for entrainment onset when different frequencies were applied. However, entrained gamma power was smaller with higher stimulation frequency, although the small sample size did not allow for a statistical analysis (110 Hz *μ* = 0.08 ±0.06 μVp; 125 Hz *μ* = 0.03 ±0.005 μVp, 145 Hz *μ* = 0.02, ±0.003 μVp).

**Figure 2.**
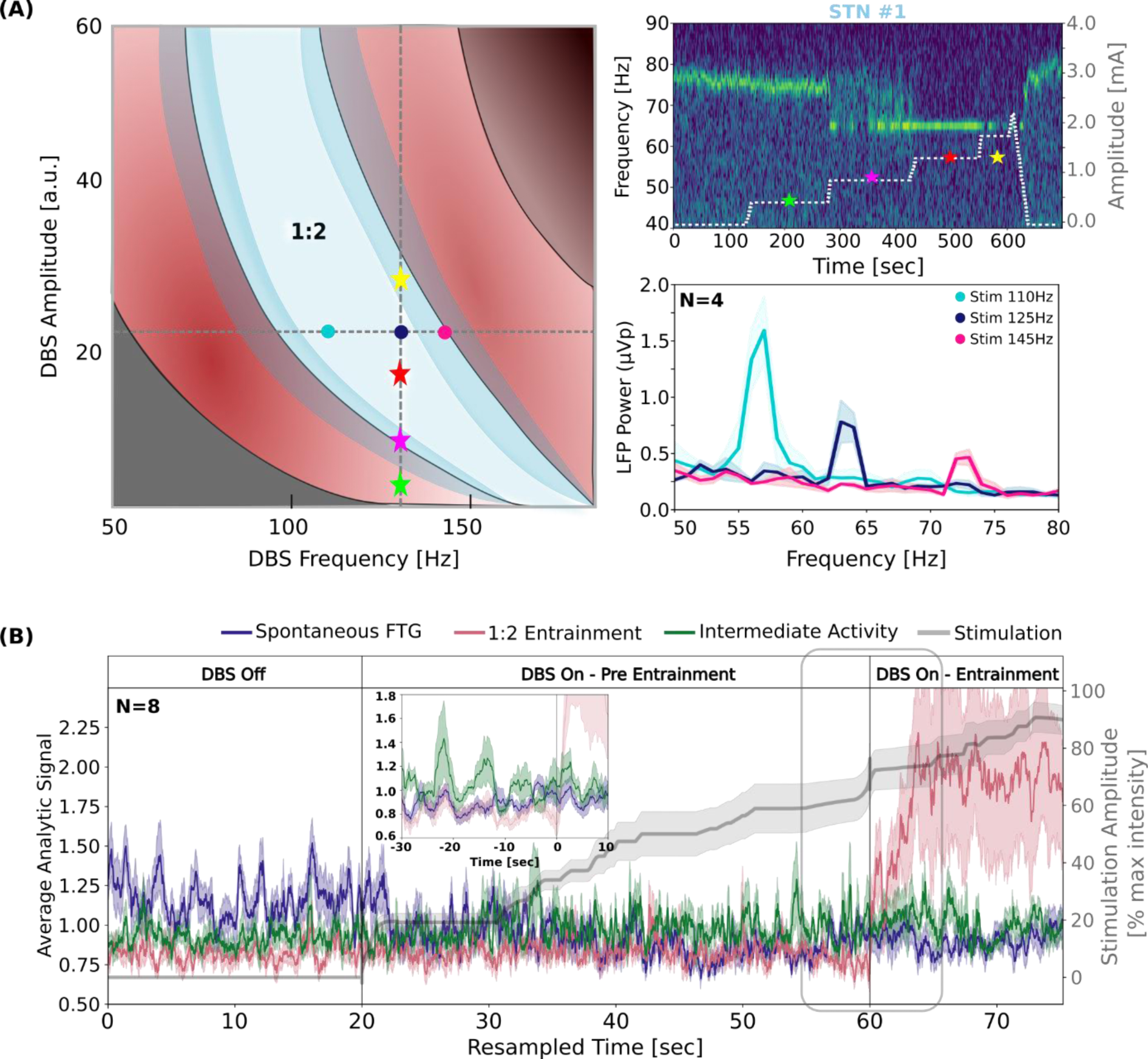
DBS-induced entrainment interacts with spontaneous FTG. **(A)** Left: Schematic representation of Arnold tongues, explaining the occurrence of 1:2 entrainment depending on deep brain stimulation (DBS) frequency and amplitude. At a stable DBS frequency (dashed vertical line indicates 130 Hz), 1:2 entrainment results from a certain range of DBS amplitudes (marked by the white area and the red star). Amplitudes in the lower or upper borders of this area (yellow and magenta stars, respectively) have a lower likelihood of causing 1:2 entrainment. The varying borders of the white area represent the dynamical nature of these borders based on neural noise. Amplitudes outside of the white area (e.g. the green star) do not cause 1:2 entrainment. This concept is depicted in the right spectrogram: Spectrogram of exemplary patient stimulated continuously at 130 Hz. The stars on the spectrogram correspond to different DBS amplitudes falling within distinct regions of the frequency-amplitude space defined by the Arnold tongue. At 1.0 mA, 1:2 entrainment is not fully present, but at 1.5 mA, it becomes noticeable, disappearing again at 2.0 mA. Similarly, at a stable DBS amplitude (dashed horizontal line in left illustration), there is a varying likelihood of 1:2 entrainment to be induced by different DBS frequencies. DBS frequency at 110, 125, and 145 Hz are denoted with turquoise, blue, and magenta dots respectively. This is shown in group power spectra of 1:2 entrainment in different DBS frequencies (right bottom plot). **(B)** Averaged analytic signal (STNs #1-8) of spontaneous FTG peak as identified in the DBS-off state (blue), averaged analytic signal of 1:2 entrainment frequency band as identified during DBS-ON (pink), and averaged power of intermediate activity (green) with increased DBS intensity (grey). All recordings were resampled in equal epochs denoted by the dashed vertical lines. Embedded figure: averaged analytic signal during real time from the eight STNs 30 seconds before the onset of entrainment. There is an increase of the intermediate activity before it entrains to ½ DBS frequency.

### 2. DBS-induced gamma entrainment interacts with spontaneous FTG

Eight patients showed spontaneous FTG in absence of DBS and allowed us to investigate the interaction between spontaneous FTG and DBS-induced entrainment in the gamma band during increasing stimulation amplitudes. **Figure 2b** shows three averaged analytic signals over time (*n* = 8 STNs): signal around their individual spontaneous FTG peaks, 1:2 entrainment frequency peaks, and the intermediate gamma range between the spontaneous FTG and the 1:2 entrainment frequencies. There is a gradual decrease of spontaneous FTG frequency shortly after switching DBS on. With higher stimulation amplitude we observed a period when gamma activity was fluctuating within the intermediate frequency range (i.e. between spontaneous FTG and entrained activity) that continues until stable entrainment frequency is reached with further increase in stimulation amplitude. This transition period is shown closely in **Figure 2b (embedded)**, revealing a tendency of increased intermediate activity before entrainment.

Entrainment power diminished and disappeared with increased DBS amplitude in one and two patients respectively. These observations follow the conceptual framework of Arnold tongues, which assumes that there is an individual region in which combinations of DBS amplitudes and frequencies will have more likelihood to induce 1:2 entrainment (**Figure 2a**).

### 3. DBS-induced gamma entrainment is modulated by movement and is linked to successful motor performance

Last, we investigated the functional relevance of DBS-induced entrainment by testing whether it is modulated by movement or influences motor performance. Nine out of 15 patients performed a finger tapping task during the DBS settings that evoked entrainment. In these patients, averaged power of the 1:2 entrainment increased during the finger tapping movements compared to rest (**Figure 3a**; Power during movement, baseline-corrected to rest *μ =* 1.41 □0.33 μVp, *P* = 0.008). Moreover, in an exemplary case we observed the presence of 1:2 entrainment only during movement (see **Figure 3b**). In the OFF DBS condition, a similar increase in spontaneous FTG power with motor performance was observed in two patients that exhibited FTG (data not shown).

**Figure 3.**
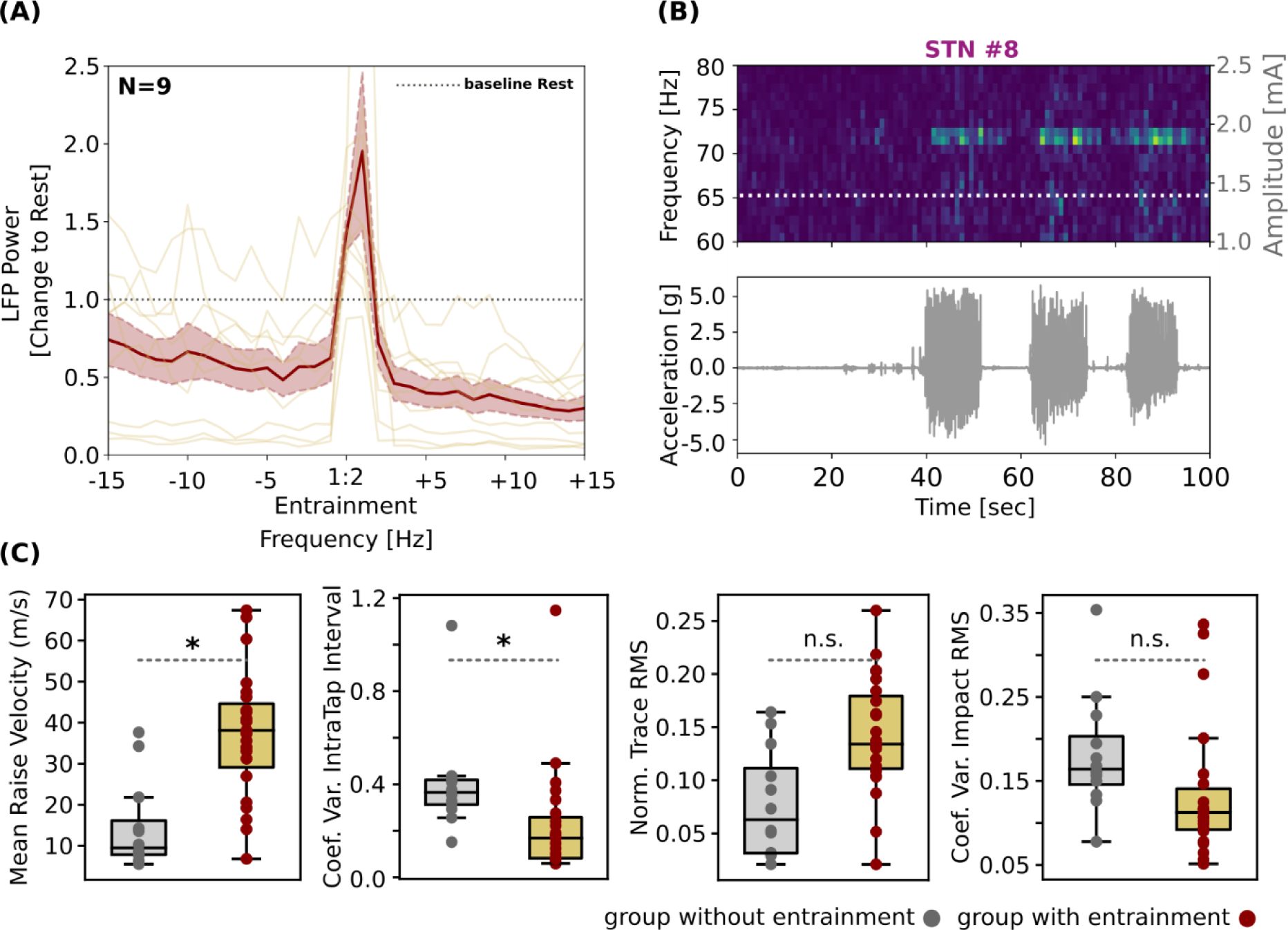
DBS-induced entrainment is modulated by movement and improves motor performance. **(A)** Mean power spectrum of 9 STNs during movement corrected to rest period. Values >1 indicate higher amplitude of entrainment during movement. **(B)** Upper plot: Spectrogram of an exemplary case of STN stimulated at 1.4 mA and 145 Hz. Bottom plot: synchronized movement traces of three blocks of finger tapping lasting 10 seconds each, with 10 seconds rest in between. Entrainment persists during the blocks of repetitive finger tapping but fades away directly after the movement is stopped. **(C)** Comparisons of the different movement metrics between the group with entrainment and the group without. It is shown that the group that exhibited entrainment performs better in repetitive finger tapping than the group that did not.

To further analyze the physiological relevance of DBS-induced 1:2 entrainment, we compared the motor performance during DBS between the group that showed entrainment (*n* = 9) and the subgroup that did not (*n* = 4). Mean raise velocity was higher (*P* = 0.017, Confidence Intervals CI: 3.705-37.59) and the coefficient of variation of intra-tap interval was lower in the entrainment group (*P* = 0.043, CI: −0.358:-0.006), indicating better tapping performance. Normalized RMS of the trace and coefficient of variation of the impact force did not yield into statistical significance (*P* = 0.058, CI: −0.007-0.123, *P* = 0.184, CI: −0.126-0.024), however all movement metrics showed a tendency towards the group with entrainment performing better. DBS amplitude had no significant effect as a fixed effect on these four metrics in LMEs (**Figure 3c**).

## Discussion

This is the first study to describe electrophysiological characteristics and functional significance of entrained gamma activity in PD patients during STN DBS. We could show that about 80% of PD patients ON medication present with DBS-induced gamma entrainment during therapeutic high-frequency DBS. We confirmed that entrainment appears with increased DBS amplitude at exactly half the frequency of the DBS frequency applied, which was irrespective of the presence of spontaneous FTG without stimulation or the occurrence of dyskinesia. If spontaneous FTG is present, we show that there is a transition period by which, with increasing DBS amplitude, the peak frequency of spontaneous FTG gradually decreases until it locks to ½ DBS frequency, which further speaks for an entrained gamma being a non-artefactual phenomenon. In the same vein, in a subset of patients the power of 1:2 entrainment is reduced with higher DBS amplitude or frequency. These observations can be interpreted within the theoretical framework of Arnold Tongues (**Figure 2**) for entrained activity. Further, our results show an enhancement of the 1:2 entrainment signal caused by short finger-tapping blocks, and patients with gamma entrainment had better motor performance as compared to those without entrainment, pointing at a prokinetic role within the motor network. Importantly, entrained gamma activity was not necessarily associated with dyskinesia. Here, we will further discuss the electrophysiological characteristics and potential functional role of DBS-induced gamma entrainment in the context of movement and DBS therapy in PD.

### DBS-induced gamma entrainment is not merely an artefact

DBS-induced 1:2 gamma entrainment has been described only recently in recordings from the STN and cortex in PD patients^9,12^ and model-based approaches have confirmed subharmonic entrainment of neural oscillations.^20,21,30^ Such subharmonic activity should be treated cautiously as a potential artefact. Several features such as medication dependency, modulation with sleep stage, and presence of dyskinesia, as well as non-linear stimulation response have been discussed supporting its physiological origin, however, its functional role is still not well understood.^30^ Here, we confirm the model predictions in a large patient group and provide further evidence that argue against the entrainment merely being an artefact of DBS. First, we showed that DBS-induced gamma entrainment interacts with the spontaneous FTG by gradually locking the FTG to half of the DBS frequency with increasing amplitudes (**Figure 2**). This observation fits the hypothesis that entrainment depends on the presence of endogenous oscillatory activity belonging to physiological processes.^11,13,31^ Second, increased DBS amplitudes led to an attenuation of the entrainment in three out of 15 cases (**Figure 1c**), a finding that confirms a non-linear response to stimulation. This attenuation of 1:2 entrainment with higher DBS intensity fits the Arnold tongue framework which suggests that entrainment only appears in certain regions of the amplitude-frequency space (**Figure 2a**).^20^ In line with this, the amplitude of entrained activity was also modulated by frequency of stimulation. Third, entrainment was modulated by movement and related to better motor performance supporting a prokinetic functional role of entrained gamma activity.

### DBS-induced gamma entrainment is modulated by movement and improves motor performance

So far, finely tuned gamma activity has been described as an oscillatory activity with a peak frequency between 60-90 Hz in the motor network in different basic ganglia nuclei,^32^ the thalamus,^33^ and motor cortex,^9^ and has been associated with level of alertness and increased motor activity.^7,33–35^ This was distinguished from a broadband gamma activation that occurs with movement and most likely reflects neuronal spiking activity over a broad frequency range (30-200 Hz). We observed an increased amplitude of gamma entrainment during movement that supports its functional relevance. Studies in different neurophysiological fields showed that entrainment requires both an endogenous oscillatory activity as well as an external stimulus. Specifically, it was shown that repetitive high-frequency stimulation enhanced local ongoing activity and that entrainment was more prominent when the stimulus was applied in frequencies closer to the intrinsic frequencies.^11,13,36^ This external manipulation of ongoing oscillatory dynamics was also found to have beneficial effects in processes such as associative learning, and to improve medical conditions, such as tinnitus.^14,18^ Entrainment caused by subthalamic DBS would typically appear in the gamma band around stimulation frequency (130 Hz) or its subharmonics (60-65 Hz). Activity in the gamma band increases with voluntary movements (40-90 Hz) and with dopaminergic medication.^5^ This increase of broadband gamma was also found to be a significant predictor of symptom alleviation, indicating its prokinetic effect. Here, we suggest that an additional increase in broad-band gamma activity during movement may result in an increase of the entrainment amplitude.

Moreover, we show that patients that exhibited DBS-induced gamma entrainment performed better in the finger tapping task than the patients that did not. This could be explained by an enhancing effect of the physiological mechanism that is related to the 1:2 entrainment frequency, in this case the prokinetic broadband gamma range. This hypothesis is supported by previous studies that showed that stimulation at 20 Hz (i.e. within the beta band of 13-35 Hz) worsened motor performance and bradykinesia.^37,38^ In this case, entrainment possibly promoted activity within the beta frequency range that is considered pathological and enhanced parkinsonian symptomatology. DBS-induced entrainment may enhance (patho)- physiological oscillatory activity, and this may explain part of the prokinetic effect of subthalamic high-frequency DBS and its effectiveness in alleviating bradykinesia. This could represent an additional, previously unexplored mechanism of DBS, indicating the potential utility of entrainment as an informative signal when adjusting DBS amplitudes for patients with Parkinson’s disease.

### Finely tuned gamma activity in the motor network and dyskinesia

FTG has been observed in Parkinson’s disease with dopaminergic medication, and in most cases it was associated with dyskinesia.^6–10^ In the framework of pro- and antikinetic activity^39^ gamma activity is considered a prokinetic brain rhythm that is observed in the ON-medication condition in parallel with improved motor performance.^40^ More recently, DBS entrained gamma band activity has been described mainly in the motor cortex in selected PD cases and used as a biomarker for dyskinesia. In this context, entrained gamma is proposed as a feedback biomarker for adaptive DBS.^41^ Our results call for a more differentiated analysis of the entrained signal. Interestingly, dyskinesia only occurred in those patients (three out of eight) that presented spontaneous FTG under dopaminergic medication before stimulation was switched on. In four out of eight patients from this subgroup, dyskinesia primarily occurred or worsened with DBS. Patients without spontaneous FTG did not show dyskinesia even though they had gamma entrainment. Nevertheless, these patients showed better motor performance as compared to those without entrained gamma pointing to its functional role within the motor network. Another interesting observation is that patients with spontaneous FTG had a better motor improvement with dopaminergic medication at the time of recording as measured by UPDRS III.

In this context one could hypothesize that dopaminergic medication induced a broader motor network facilitation in this cohort that is represented by spontaneous FTG leading the network towards a state that is prone to dyskinesia. Entrained gamma activity may lead to a similar but more constrained motor network activation that is less likely to overshoot resulting in dyskinesia. Only if the network is already at an elevated level of activity, dyskinesia may break through. In line with this, DBS induced gamma entrainment is less likely to occur in the OFF dopaminergic medication condition.^12^ Depending on the prokinetic activation level of the motor system, which is modulated by dopamine, focal network stimulation (STN DBS) can lead to gamma entrainment with improved motor performance. This could potentially result in dyskinesia (high level of activation represented by spontaneous FTG) or not (intermediate level), when the gamma activity increase is more rigid confined within the borders of the subharmonic DBS frequency. One could hypothesize that DBS partially works by motor network activation confined to a narrow-band gamma frequency that may be less propagated across the network, but is balanced by the underlying dopaminergic network state activation. Reduction in dopaminergic medication with STN DBS is a prerequisite in clinical routine to allow effective stimulation without adverse motor and non-motor effects in PD patients. The specific interplay between broad network state changes induced by dopaminergic medication, and more focal and narrow-band stimulation induced activation pattern need to be characterized in future studies.

### DBS-induced gamma entrainment and closed-loop DBS

DBS-induced gamma entrainment has been recently introduced as a feedback signal for closed-loop DBS.^41^ Here, stimulation-entrained gamma is used to adjust DBS parameter, specifically to control DBS amplitude reduction because this biomarker has been related to dyskinesia. Entrained gamma on one hand is a potential robust closed-loop DBS input signal as it has an entirely predictable frequency and large peak amplitude, which can be also used in chronic devices with limited spectral recording bandwidth. Our data, on the other hand, suggest that this biomarker is not solely associated with dyskinesia but could predict a prokinetic state that should be maintained. Thus, linking the entrained gamma signal to reduction in stimulation amplitude may lead to reduced effectiveness of DBS in a state of reduced dopaminergic stimulation. The positive trend between gamma entrainment and motor performance we observed in our cohort would support to maintain DBS during gamma entrainment if dyskinesia is absent. This contradicts the results from three patients in Oehrn *et al.*^41^ that reported that entrained FTG can predict hyperkinetic states in PD patients at home and was successfully applied to reduce residual motor fluctuations. A deeper understanding of the impact of dopaminergic medication, daily activities, and dyskinesia severity on DBS gamma entrainment is required. So far, this variability poses an ongoing challenge for the utility of gamma entrainment in closed-loop DBS algorithms.

### Limitations

We also have to consider some limitations of the current study. First, the patients were recorded under two protocols, which had an impact on the selection of the recording contacts. In patients with 3389 electrodes the contact pairs with the highest beta band activity were used for recording, while in patients with SenSight electrodes contact selection for stimulation was based on gamma entrainment during a previous systematic survey. This precluded further analysis of contact specificity for gamma entrainment but unlikely had a significant impact on the reported results. Second, the stimulation protocol was focused on short-term modulation of neuronal signals by stimulation and its clinical consequences for motor performance. This precluded the assessment of long-term stimulation effects especially on dyskinesia that sometimes occur only after minutes or hours of high amplitude stimulation. Third, the patient group that did not show gamma entrainment was limited to four patients, which led to an imbalance in power for the comparison between groups. Nevertheless the results were consistent across the cohort despite the small sample size, and are supported by previous reports, where enhanced gamma activity is linked to improved motor performance.

## Conclusion

Here, we propose a functional role of DBS-induced gamma entrainment in the STN of patients with Parkinson’s disease for motor improvement. We show that the amplitude of the entrainment increased during movement and that entrainment is not necessarily related to dyskinesia. We also show that entrainment interacts with ongoing oscillatory activity, DBS intensity and frequency of stimulation, an observation that is well interpreted by the concept of Arnold tongues. These findings support a prokinetic role of DBS-induced gamma entrainment for PD that may lead to motor improvement with DBS.

## Acknowledgments

We would like to thank the patients that participated in the study. We would like to thank Ulrike Uhlig and Rose Franx for their help with organizing and assisting during the recordings.

