## Supplemental Figure 1 for "DBS-induced gamma entrainment as a new biomarker for motor improvement with neuromodulation"

### SUPPLEMENTARY MATERIAL

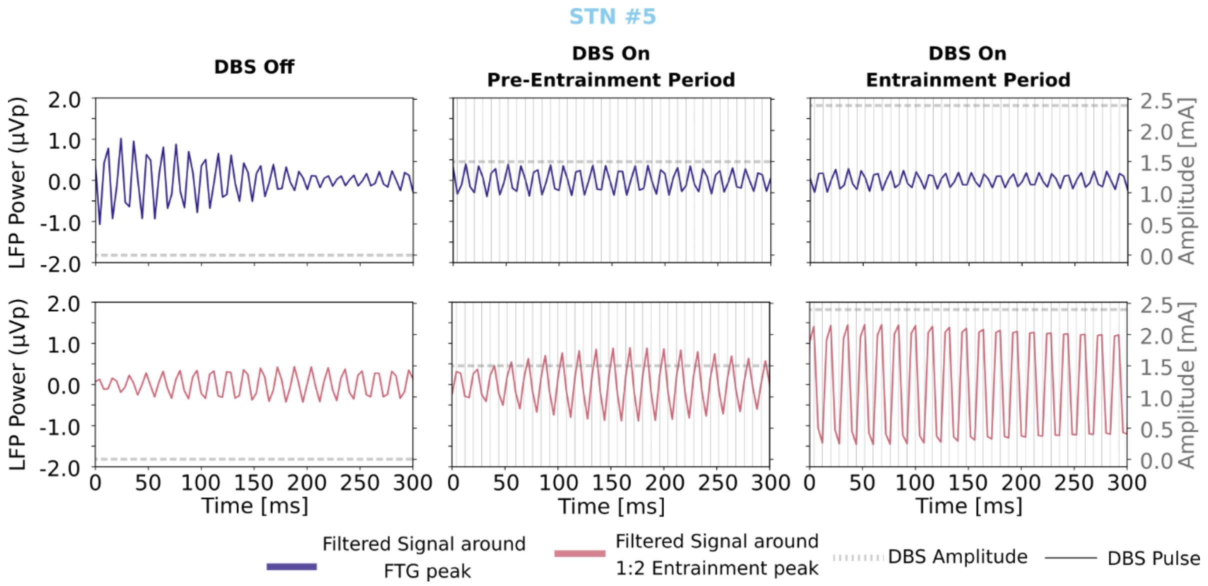


**Supplementary Figure 1. Gamma entrainment in an example LFP recording.** 300 ms filtered 5 Hz around FTG peak (76 Hz in purple) and around the entrainment peak (63 Hz in pink) during DBS-off, pre-entrainment period (DBA amplitude: 1.5 mA), and during highest DBS amplitude where entrainment was observed (2.5 mA). DBS was applied with a frequency of 125 Hz and pulse width of 60 µs, therefore estimated DBS pulses are plotted every 8 ms. During 1:2 entrainment it is visible that a peak of activity appears every two DBS pulses.
